# A Set of Diagnostic Tests for Detection of Active *Babesia duncani* Infection

**DOI:** 10.1101/2024.03.25.24304816

**Authors:** Meenal Chand, Pratap Vydyam, Anasuya C. Pal, Jose Thekkiniath, Dounia Darif, Zeng Li, Jae-Yeon Choi, Ruben Magni, Alessandra Luchini, Laura Tonnetti, Elizabeth J Horn, Danielle M Tufts, Choukri Ben Mamoun

## Abstract

Human babesiosis is a rapidly emerging and potentially fatal tick-borne disease caused by intraerythrocytic apicomplexan parasites of the *Babesia* genus. Among the various species of *Babesia* that infect humans, *B. duncani* has been found to cause severe and life-threatening infections. Detection of active *B. duncani* infection is critical for accurate diagnosis and effective management of the disease. While molecular assays for the detection of *B. duncani* infection in blood are available, a reliable strategy to detect biomarkers of active infection has not yet been developed. Here, we report the development of the first *B. duncani* antigen capture assays that rely on the detection of two *B. duncani*-exported immunodominant antigens, BdV234 and BdV38. The assays were validated using blood samples from cultured parasites in human erythrocytes and *B. duncani*-infected laboratory mice at different parasitemia levels and following therapy. The assays display high specificity with no cross-reactivity with *B. microti*, *B. divergens*, *Babesia* MO1, or *P. falciparum.* The assay also demonstrates high sensitivity, detecting as low as 115 infected erythrocytes/µl of blood. Screening of 1,731 blood samples from diverse biorepositories, including previously identified Lyme and/or *B. microti* positive human samples and new specimens from field mice, showed no evidence of *B. duncani* infection in these samples. The assays could be useful in diverse diagnostic scenarios, including point-of-care testing for early *B. duncani* infection detection in patients, field tests for screening reservoir hosts, and high-throughput screening such as blood collected for transfusion.

**Short summary:** We developed two ELISA-based assays, BdACA38 and BdACA234, for detecting *B. duncani*, a potentially fatal tick-borne parasite causing human babesiosis. The assays target two immunodominant antigens, BdV234 and BdV38, demonstrating high specificity (no cross-reactivity with other *Babesia* species or *Plasmodium falciparum*) and sensitivity (detecting as low as 115 infected erythrocytes/µl). The assays were validated using in vitro-cultured parasites and infected mice. Screening diverse blood samples showed no evidence of *B. duncani* active infection among 1,731 human and field mice blood samples collected from the north-eastern, midwestern, and western US. These assays offer potential in diverse diagnostic scenarios, including early patient detection, reservoir animal screening, and transfusion-transmitted babesiosis prevention.

## INTRODUCTION

In recent years, vector-borne infectious diseases have emerged as a significant global public health concern (1). The expansion of the geographic range of the tick vectors has led to a notable increase in tick-borne diseases (TBDs), which now account for more than 75% of all reported vector-borne diseases in the United States annually (2–4). Among these tick-borne diseases is human babesiosis, an emerging infectious disease caused by intraerythrocytic apicomplexan parasites of the *Babesia* genus (5). In 2011, the disease became a US national notifiable condition in 18 states, which led to a significant increase in the number of reported cases. A recent report by the US Centers for Disease Control and Prevention (CDC) documented a rapid surge in human babesiosis cases over the past decade, 16,456 clinical cases were reported between 2011 and 2019 primarily in 37 states, with 98.2% of cases reported concentrated in 10 states (6). While ticks are the main vehicle of transmission of *Babesia* parasites, blood transfusion remains a major concern despite screening efforts.

To date, eight species of *Babesia* have been identified as human pathogens and documented to cause mild to severe babesiosis with some cases resulting in fatal outcomes (7). The majority of clinical cases reported to the CDC are attributed to *Babesia microti* (8, 9), whereas only a few cases of *B. duncani* babesiosis have been documented in the United States, primarily in Washington, Oregon, and California. However, it is crucial to acknowledge that some of the *B. microti* cases might be attributed to *B. duncani.* This observation underscores the significance and justifies the necessity for a new and highly sensitive diagnostic test capable of distinguishing between the two Babesia species. Indeed, as per the CDC Babesiosis Surveillance Report (2011–2015) in the United States, three babesiosis cases were linked to *B. duncani* in Maryland and Connecticut (10) based on serological criteria. This underscores the concern regarding the presence and potential threat of *B. duncani* on the East Coast and validates the need for a new, highly sensitive diagnostic test that is specific to *B. duncani*.

In confirmed cases of *B. duncani* babesiosis, the parasite was found to induce fulminant and often fatal infections in both immunocompetent and immunocompromised individuals (11–14). Available data suggest that the hard tick *Dermacentor albopictus* serves as the vector for *B. duncani* (13).

To date, the true incidence of *B. duncani* babesiosis remains unknown. Accurate determination of the true prevalence of *B. duncani* babesiosis is challenging due to the absence of specific and reliable diagnostic tests that can determine active infection. Over the years, various diagnostic techniques including microscopy, serological tests like immunofluorescence assay (IFA), immunoblot and enzyme-linked immunosorbent assay (ELISA), and nucleic acid-based tests such as nested PCR, reverse transcription-polymerase chain reaction (RT-PCR), and transcription-mediated amplification (TMA), have been employed to detect *Babesia* infections. Morphologically, *B. duncani*-infected red blood cells (RBCs) are indistinguishable from those infected with *B. microti* or other *Babesia* species and bear resemblance to the ring-stage forms of *Plasmodium falciparum*, the causative agent of severe human malaria. Therefore, microscopy cannot be reliably used to diagnose *B. duncani* active infection in a blood sample. With the exception of cases confirmed by both microscopy and PCR (14–19), most unverified cases attributed to this parasite have relied on serological assays, which cannot reliably distinguish between past and active infections. A 2018 publication by Scott and Scott summarized reports by physicians and naturopathic physicians of 1119 cases of *B. duncani* between 2011 and 2017 in Canada, with most cases occurring in the Pacific Coast region (20). However, no positive blood smears or details about the sensitivity or specificity of the serological and molecular assays used were described in that report. Furthermore, no appropriate controls were included in these studies.

Therefore, these claims warrant further validation, which could be facilitated by sharing blood samples with established biorepositories and large-scale blood screening in different geographic areas in the US. Nucleic acid-based detection techniques are indeed more specific and sensitive and have been effective in identifying positive *B. microti* blood samples. Nevertheless, in some instances, these techniques have been shown to detect residual parasite DNA or RNA even after the parasite has been eliminated (21, 22). Recent studies using microscopic examination of blood samples from *B. microti*-infected animals treated with drugs that cleared the infection demonstrated the presence of residual DNA after treatment with antibabesial drugs (22). Studies using *B. microti*-infected blood revealed that antigen detection assays, which rely on the detection of highly abundant and often immunodominant secreted antigens of the parasite, exhibit a stronger correlation with active *B. microti* infection compared to Nuclic Acid-detection assays (23). Therefore, efforts to develop similar assays for the detection of active infections by other *Babesia* species could be an invaluable resource to estimate the true incidence of babesiosis caused by each of the *Babesia* species that cause infection in humans. This is particularly important in light of the rapid environmental changes impacting the transmission of these diseases and other TBDs (24, 25).

A significant breakthrough in the field of *B. duncani* biology emerged with the establishment of the first continuous *in vitro* culture system of *B. duncani* in human erythrocytes (26–28). Furthermore, the recent sequencing and annotation of the parasite’s genome (29) made it possible to apply a multi-faceted approach that incorporates genomic, transcriptomic, and proteomic analyses to identify unique and species-specific proteins that are exported by the parasite from infected RBCs into the host plasma.

In this study, we report the discovery of two highly reliable biomarkers for active *B. duncani* infection. *Babesia duncani-*specific antigen capture assays (BdACAs) that detect these antigens were developed and exhibit high sensitivity, are capable of detecting as low as 115 parasites/µl of blood and can distinguish active infections from past exposures, and can distinguish between *B. microti* and *B. duncani* to provide accurate species infections, important for public health.

## MATERIALS AND METHODS

### Ethics statement

All animal experiments described in this study followed the Yale University institution guidelines for the care and use of laboratory animals and were approved by the Institutional Animal Care and Use Committee at Yale University (protocol number 2023-07689).

### *Babesia duncani* infection in mice

C3H/HeJ or SCID mice (C.B-17/IcrHsd-Prkdcscid) 5-6 week, female mice from Jackson laboratories were infected by injecting 1**×**10^6^ or 1x 10 ^7^ *B. duncani* WA-1 strain or *B. microti* LabS1 strain, respectively by infected erythrocytes intravenously (IV) from the stock mouse. Blood samples were collected at different time points by a retro-orbital bleed or cardiac puncture, and parasitemia was monitored by light microscopy of Giemsa-stained blood smears. Where indicated, plasma or serum was separated from whole blood by centrifugation at 6000g for 10 min, and packed mouse RBC (mRBC) was lysed with 1% saponin. Hemolysate and parasite pellets were collected after centrifugation at 9300g for 10 min at 4°C.

### Babesia duncani in vitro culture

*Babesia duncani* WA-1 parasites were cultured continuously in human erythrocytes in DMEM-F12 media (28), as described previously (26). (Included in Supplementary methods).

### Isolation of extracellular vesicles from plasma, culture supernatant, and hemolysate

Extracellular membrane vesicles containing protein cargo released from the parasite were isolated from supernatant/plasma and hemolysate of *B. duncani in vitro* and *in vivo* culture by EXO Quick ULTRA EV isolation kit (EQULTRA-20A-1) following the user manual. Where indicated, vesicles were also isolated by the ultracentrifugation method, as mentioned previously (30). Briefly, 1.5 ml of human RBC (hRBC) and mRBC samples (uninfected and *B. duncani* infected plasma/serum or supernatant and hemolysate) were sequentially centrifuged at 500g for 30 min, followed by 16,000g for 45 min. These samples were then centrifuged at 120,000g for 14h at 4°C using a Sorvall MTX 150 micro-ultracentrifuge with an S52-ST swinging bucket rotor (Thermo Fisher) and ultracentrifuge pellet (Up/UHp) and ultracentrifugation supernatant (Us/UHs) fractions were collected (31).

### Cloning, expression, and purification of recombinant proteins

Plasmid DNA constructs for BdV234 and BdV38 were synthesized by GenScript, Inc., NJ. Details description of protein expression and purification provided in the supplementary methods.

### Immunodetection of BdV234 and BdV38

Polyclonal antibodies raised in rabbits (Cocalico Biologicals, Inc., PA) against the indicated *B. duncani* proteins were purified and used for the immunodetection analysis as described in detail (32). Briefly, supernatant (S), hemolysate (H), and parasite pellet (P) fractions collected either from *B. duncani in vitro* culture in hRBC (18% parasitemia) or *B. duncani* infected (24% parasitemia) or uninfected mRBCs were mixed with 4X Laemmli sample buffer (3:1 ratio), boiled at 80°C for 5 min and separated on 4-20% Mini-protean gels (Bio-Rad, P4568093). The gels were then transferred onto nitrocellulose membranes (Bio-Rad, 1620115), probed with anti-BdV234 and BdV38 primary antibodies (1:250 dilution) and goat-anti-rabbit horseradish peroxidase (HRP)-conjugated IgG Ab (1: 5000 dilution, (Thermo Fisher, 31466). Blots were developed by Super signal^TM^ West Pico PLUS chemiluminescent substrate (Thermo Fisher, 34577) and imaged using the LI-COR Odyssey-Fc imaging system.

### Immunofluorescence assay (IFA)

Immunofluorescence assay was performed as reported previously (30, 32) and described briefly in the supplementary methods.

### BdACA using *B. duncani in vitro* culture supernatant and hemolysate fractions

*Babesia duncani* antigen capture sandwich ELISAs were performed as described earlier (22, 23), with few modifications. Briefly, 96-well plates were coated with 5µg/ml of capture Abs (BdV234 and BdV38) and incubated for 2h at room temperature, followed by blocking with 5% BSA in PBST (0.05% Tween-20) at 37°C for 1h. Plates were then incubated overnight at 4°C with either (1) BdV38 and BdV234 recombinant protein (100 ng/well), (2) a 1 in 10 dilution of *B. duncani in vitro* culture supernatant, hemolysate, and whole blood, or (3) plasma/serum (heat-inactivated for 30 min at 56°C), hemolysate and whole blood from uninfected or *B. duncani* infected C3H/HeJ mice. Whole blood samples from American Red Cross (ARC), Lyme Disease Biobank (LDB), and field-derived mouse samples were incubated similarly. The plates were then washed four times with PBST, and 5 μg/ml biotinylated detection Ab (BdV234 and BdV38) were added and incubated at room temperature for 2h. Following this, plates were washed four times with PBST and incubated with HRP conjugated streptavidin antibody in 1:5000 dilution at room temperature for 1h, followed by washing and incubation with 50 μl 3,3’,5,5’ tetramethylbenzidine (TMB) substrate for 7 min in the dark. The reaction was stopped by adding 50 μl of 0.1N HCl, and the Optical Density at 450 nm was measured using a BioTek Synergy Mx plate reader.

### Detection of BdV234 and BdV38 antigens in hemolysate from drug-treated *B. duncani in vitro* culture or infected mouse blood

*In vitro B. duncani* culture with an initial parasitemia of 1% (cultured in 5% hematocrit with DMEMF-12 containing hRBCs) was exposed to tafenoquine (SML0396, Sigma Aldrich) at 1x IC_50_ (3 µM) and 2x IC_50_ (6 µM) concentrations for three generations. Both untreated and tafenoquine treated parasite cultures were collected at specified time points (0, 3, 6, 12, 24, 36, 48, 60, and 72 hours post treatment), lysed with 1% saponin, and the resulting hemolysates were used for ELISA. Untreated infected and uninfected hRBCs were included as controls. Similarly, hemolysates were prepared from blood from 12 C3H/HeJ (3 male, 3 female) mice infected with a high dose of *B. duncani* (10^6^ parasites) and used in ELISA to detect BdV234 and BdV38. Six of these mice were treated with tafenoquine for 5 days (DPI 3-7), and the remaining six were treated with vehicles (PEG400). Vehicle-treated and uninfected mice hemolysates were used as controls.

### Genomic DNA extraction and Real-Time PCR

To further confirm the presence or absence of *B. duncani* DNA in infected blood, qPCR analyses were conducted as previously reported (33). Briefly, blood from *B. duncani in-vitro* culture and *B. duncani* infected mouse at various parasitemia levels were collected at specific time points post-infection, followed by gDNA isolation by the DNeasy Blood and Tissue kit (Qiagen, 69504). The isolated genomic DNA were used to amplify the *B. duncani* internal transcribed spacer (ITS) sequence in its nuclear rRNA (18s-rRNA) by using BdITS1-F 5′-GCTTCCTAACCCGAGACCAA-3′ and BdITS1-R 5′-CACTGGCGGGGTGAAAAGTA-3′ primers (34). The reaction mixture contained 1× advanced universal SYBR green super mix (1725270; Bio-Rad), 1 μl of each primer (10 μM), and 2 μl of template DNA.

### Collection of biological samples used for screening *B. duncani* antigen/infection

Two hundred human whole blood samples were obtained from the American Red Cross (ARC), of which 100 samples were positive for *B. microti* infection as detected by transcriptionally mediated amplification (TMA) positive and the remaining 100 samples were TMA negative for *B. microti*, the samples were collected in areas endemic for *B. microti.* (Connecticut, Delaware, Massachusetts, Maryland, Maine, Minnesota, New Hampshire, New Jersey, New York, Pennsylvania, Virginia, Vermont). Additionally, 770 whole blood samples were obtained from the Lyme Disease Biobank (LDB) with 317 samples enrolled with signs and symptoms of early Lyme disease and 282 controls as previously described (35) and were collected from East Hampton, Martha’s Vineyard, and Wisconsin (2014–2020). Similarly 171 samples were collected from the west coast (California), including 11 enrolled with early Lyme disease, 10 controls, and 150 with peresistent Lyme symptoms. Whole blood samples were also obtained from 761 white-footed mice (*Peromyscus leucopus*), a prominent reservoir host of *B. microti*, (unscreened) that were collected from western Pennsylvania, USA, and used in this study.

## RESULTS

### Identification of *B. duncani* secreted antigens

The genome of *B. duncani* has been extensively studied and characterized. Its annotation identified 4222 proteins, of which 479 are predicted to be members of the parasite’s secretome (29). To identify proteins exported by the parasite into the host red blood cell and/or host environment, we used a Nanotrap-based proteomics approach (36, 37) on hemolysate (H) and secreted (S) fractions collected from a culture of *B. duncani*-infected human erythrocytes as shown in **Fig. 1A**. Mass spectrometry analysis identified 27 proteins in the S fraction (**Table 1**) and 39 proteins in the H fraction. Of these exported proteins, BdV234 and BdV38 were selected based on their peptide abundance, high expression levels of their encoding genes as determined by RNAseq, and predicted antigenicity profiles (**Table 1**). The genes encoding these two proteins were cloned into the pGEX 6p-1 and pET21a (+) expression vectors, respectively, and the encoded recombinant proteins were expressed in *E. coli,* purified on glutathione Sepharose high-performance (BdV234) and Ni-NTA agarose (BdV38) affinity columns and used to generate specific polyclonal antibodies in rabbits. The antibodies were then affinity-purified and evaluated for their specificity using recombinant proteins as well as total extracts from a culture of *B. duncani*-infected erythrocytes. The resulting affinity-purified anti-BdV234 and anti-BdV38 antibodies were used to detect the expression and cellular distribution of the native proteins in the supernatant (S), hemolysate (H), or pellet (P) fractions from cultures of *B. duncani*-infected human erythrocytes and control uninfected human erythrocytes (**Fig. 1B**). Consistent with the export of these proteins, immunoblot analyses identified the native BdV234 and BdV38 in all three fractions while no signal could be detected in the fractions isolated from the uninfected erythrocytes **(Fig. 1B)**. We then assessed the cellular distribution of the same antigens in mice infected with *B. duncani*. Interestingly, whereas the antigens could be readily detected in the H and P fractions prepared from mouse blood collected from *B. duncani*-infected C3H/HeJ mice, the signal from the plasma (Pl) fraction was very weak, likely the result of dilution in the large volume of plasma collected from infected animals **(Fig. 1B**). Additionally no signal could be detected from the Pl, H, or P fractions prepared from blood collected from uninfected mice, further confirming the specificity of the antibodies raised against these proteins (**Fig. 1B**).

**Figure 1.**
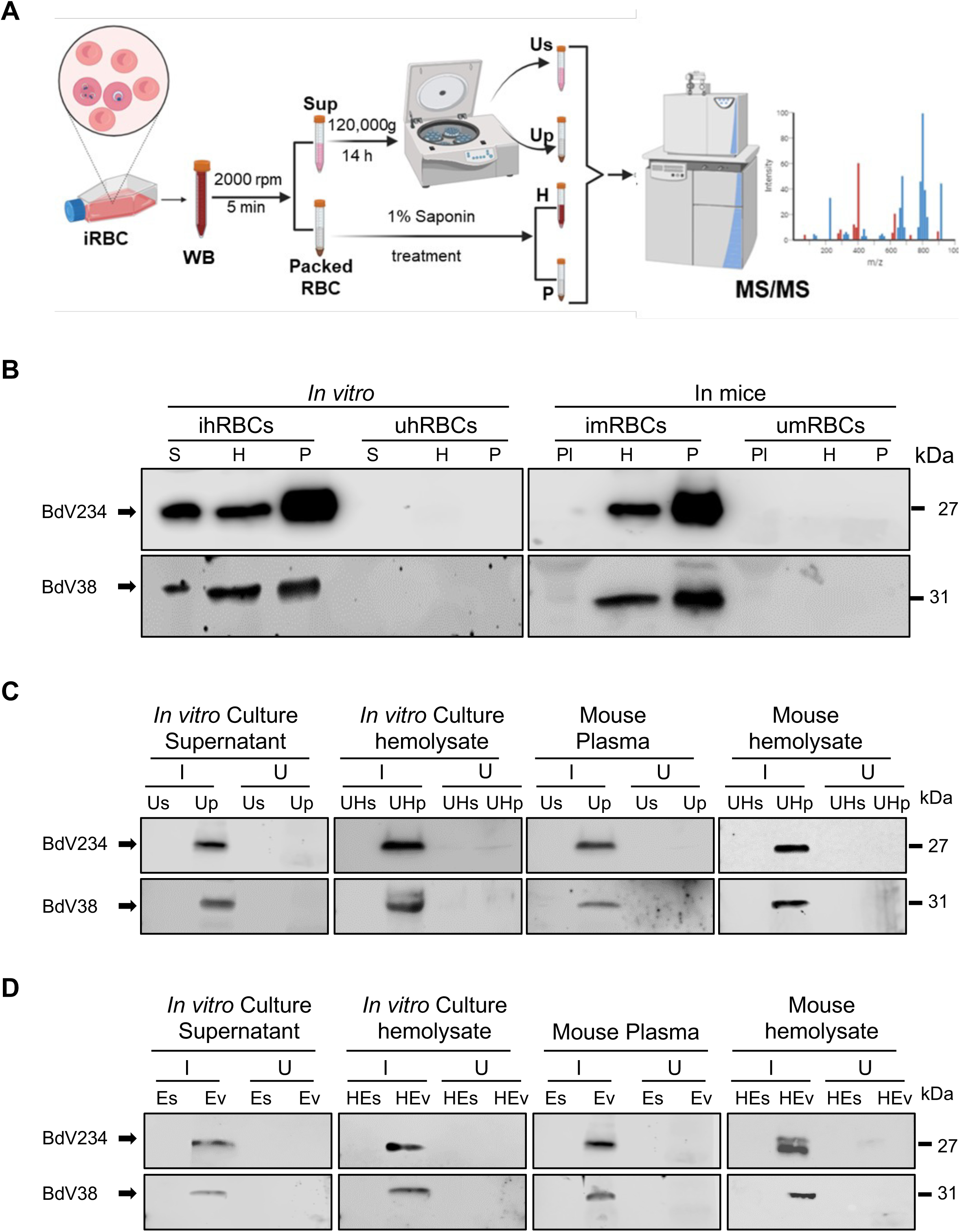
Identification and detection of *B. duncani* secreted antigens. **(A)** Schematic presentation for qualitative analysis of *B. duncani* secretome by nano-trap-based proteomics approach (NTPA) utilizing ultracentrifugation and a tandem mass spectrometry analysis (MS/MS) to identify *Babesia duncani* secreted proteins. (**B)** Immuno detection of secreted BdV234 and BdV38 antigens from *B. duncani* infected human *in vitro* and mouse (ihRBCs, imRBCs) culture supernatant (S), plasma (Pl), hemolysate (H) and parasite pellet (P) and uninfected RBC (uhRBC, umRBC) with their corresponding protein size. (**C)** Western blot analysis to detect BdV234 and BdV38 secreted antigens from extracellular vesicular fractions obtained by ultracentrifugation approach using *B. duncani*-infected human and mouse RBCs. After ultracentrifugation, the soluble fraction Us and the pellet fraction Up were collected from the culture supernatant/plasma, and UHs and UHp fractions were obtained from the hemolysate. Our data indicated that these proteins containing extracellular vesicles accumulate in the pellet fraction Up/UHp from *in vitro* as well as from *in vivo* supernatant/plasma or hemolysate. These measurements were conducted under both infected (I) and uninfected (U) conditions. **(D)** Detection of BdV234 and BdV38 proteins from extracellular vesicle supernatant (Es), and vesicle-rich pellet (Ev) fractions from both *B. duncani* infected and uninfected *in vitro* culture supernatant and mouse plasma. Similar, detection of protein from extracellular vesicles (EHs) and pellet (EHv) fractions of both human and mouse hemolysate samples.

**Table 1.** List of proteins identified in the supernatant of *B. duncani*-infected human erythrocytes using nano trap technology.

| Accession ID | Description | Protein FDR Confidence | Number of Peptides | Number of amino acids | MW [kDa] | RNAseq (TPM)# |
| --- | --- | --- | --- | --- | --- | --- |
| BdWA1_000818-T1 | Actin, putative | High | 5 | 376 | 41.9 | 6836.72 |
| BdWA1_000589-T1 | phosphatidylinositol-4-phosphate 5-kinase, putative | High | 4 | 1074 | 121.8 | 278.21 |
| BdWA1_002814-T1 | Thioredoxin domain\|Thioredoxin, conserved site\|Thioredoxin-like superfamily | High | 4 | 490 | 54.4 | 1398.21 |
| BdWA1_000266-T1 | hypothetical protein, conserved | High | 3 | 498 | 57.4 | 21.68 |
| BdWA1_001049-T1 | Hsp20/alpha-crystallin family, putative | High | 3 | 177 | 20.1 | 8294.36 |
| BdWA1_002869-T1 | hypothetical protein, conserved | High | 3 | 215 | 24.6 | 778.09 |
| BdWA1_003373-T1 | hypothetical protein | High | 1 | 127 | 12.5 | 8199.71 |
| BdWA1_001726-T1 | RAP domain/Core histone H2A/H2B/H3/H4, putative | High | 3 | 132 | 14.5 | 26809.25 |
| **BdWA1_002223-T1** | **BdV234**<br>(GPI-anchored protein BdGPI8) | **High** | **2** | **242** | **27.2** | **1655.60** |
| BdWA1_001531-T1 | hypothetical protein | High | 3 | 375 | 41.4 | 4774.64 |
| BdWA1_003044-T1 | heat shock protein 70 precursor, putative | High | 2 | 644 | 70.6 | 2621.88 |
| **BdWA1_001730-T1** | **BdV38**<br>(putative EF-hand domain) | **High** | **1** | **292** | **33** | **1426.92** |
| BdWA1_001385-T1 | 40S ribosomal protein S3 protein, putative | High | 1 | 224 | 25 | 1702.62 |
| BdWA1_000947-T1 | heat shock protein 90 | High | 2 | 713 | 82.3 | 4798.62 |
| BdWA1_001677-T1 | Sodium: dicarboxylate symporter family, putative | High | 1 | 789 | 89.4 | 272.46 |
| BdWA1_002995-T1 | NAC domain-containing protein, putative | High | 1 | 159 | 17.1 | 575.40 |
| BdWA1_001746-T1 | hypothetical protein, conserved | High | 1 | 374 | 42.8 | 667.65 |
| BdWA1_001707-T1 | heat shock protein 70 precursor, putative | High | 2 | 654 | 71.9 | 0.98 |
| BdWA1_001359-T1 | histone H3, putative | High | 2 | 136 | 15.4 | 21963.75 |
| BdWA1_002631-T1 | Porin domain superfamily | High | 1 | 298 | 32.9 | 572.95 |
| BdWA1_000088-T1 | hypothetical protein | High | 1 | 601 | 68.4 | 17.86 |
| BdWA1_001694-T1 | Enolase,N-terminal domain / Enolase, C-terminal TIM barrel domain-containing protein, putative | Medium | 1 | 442 | 47.6 | 4.71 |
| BdWA1_000240-T1 | BdGPI17 | Medium | 1 | 618 | 69.8 | 1674.59 |
| BdWA1_000444-T1 | myosin A | Medium | 1 | 827 | 92.4 | 7.04 |
| BdWA1_002345-T1 | cytochrome c oxidase subunit 2, putative | Medium | 1 | 168 | 19.1 | 353.85 |
| BdWA1_002720-T1 | transporter, putative | Medium | 1 | 445 | 50.6 | 1769.55 |
| BdWA1_000077-T1 | Armadillo-type fold Armadillo-like helical\|Armadillo repeat-containing protein 6\|Armadillo | Medium | 1 | 2574 | 283.2 | 947.97 |

\* FDR (False discovery rate)

### Evidence of vesicular-mediated secretion of BdV234 and BdV38 from *B. duncani*-infected erythrocytes

Recent studies in *B. microti* and *B. divergens* have demonstrated that several proteins of these parasites’ are actively exported by using vesicular-mediated export mechanisms (30, 38). To assess the mode of export of BdV234 and BdV38, the culture S and H fractions of *B. duncani-*infected human erythrocytes, as well as the Pl and H fractions of *B. duncani*-infected mice were subjected to ultracentrifugation at 120,000 x g for 14h at 4°C to separate the soluble (Us/UHs) and vesicle-associated membrane (Up/UHp) fractions. These resulting fractions were then analyzed by immunoblotting using anti-BdV234 and anti-BdV38 antibodies to assess the presence or absence of the antigens. As shown in **Fig. 1C**, the proteins were primarily found associated with the Up/UHp fractions in both human and mouse infected samples (**Fig. S1-A**), indicating that once exported from the infected erythrocytes, these antigens are associated with vesicles. This association was further validated using the ExoQuick® vesicle isolation system, which was used to separate the supernatant fraction (Es/EHs) from the vesicle-rich pellet fraction (Ev/EHv) on plasma, supernatant, and hemolysate fractions collected from uninfected or *B. duncani*-infected blood (**Fig. S1-B**). As shown in **Fig. 1D**, both antigens were detected in the vesicle fractions (E_V_ or HE_V_) from the S, H, and Pl fractions obtained from *B. duncani*-infected human and mouse RBCs, respectively. As a control, no signal could be detected in similar fractions prepared from blood collected from uninfected human erythrocytes *in vitro* or uninfected C3H/HeJ mice (**Fig. 1D**). Together these data demonstrate that BdV234 and BdV38 are exported by *B. duncani*-infected erythrocytes through a vesicular-mediated mechanism.

### Cellular localization of selected *B. duncani* secreted proteins

In order to investigate the localization and cellular distribution of BdV234 and BdV38 secretory proteins in *B. duncani*, immunofluorescence assays (IFA) were conducted on uninfected or *B. duncani*-infected human erythrocytes by using affinity-purified BdV234 and BdV38 antibodies from rabbit sera. Anti-rabbit Alexafluor-488 was used for the detection of BdV234 and BdV38, and α-Band III antibody was used as a membrane marker for human RBCs. Interestingly, BdV234 was not only found to be localized inside the parasites but was also found in the host erythrocytes displaying punctate vesicular staining (**Fig. 2**), which indicating that this GPI-anchored protein is secreted by the parasite into the host cytosol. On the other hand, BdV38 was found to be localized primarily within the parasites. The rabbit pre-immune sera served as a control in our IFA, and no localization signal with BdV234 and BdV38 was observed other than the nucleus and RBC membrane (**Fig. 2**).

**Figure 2.**
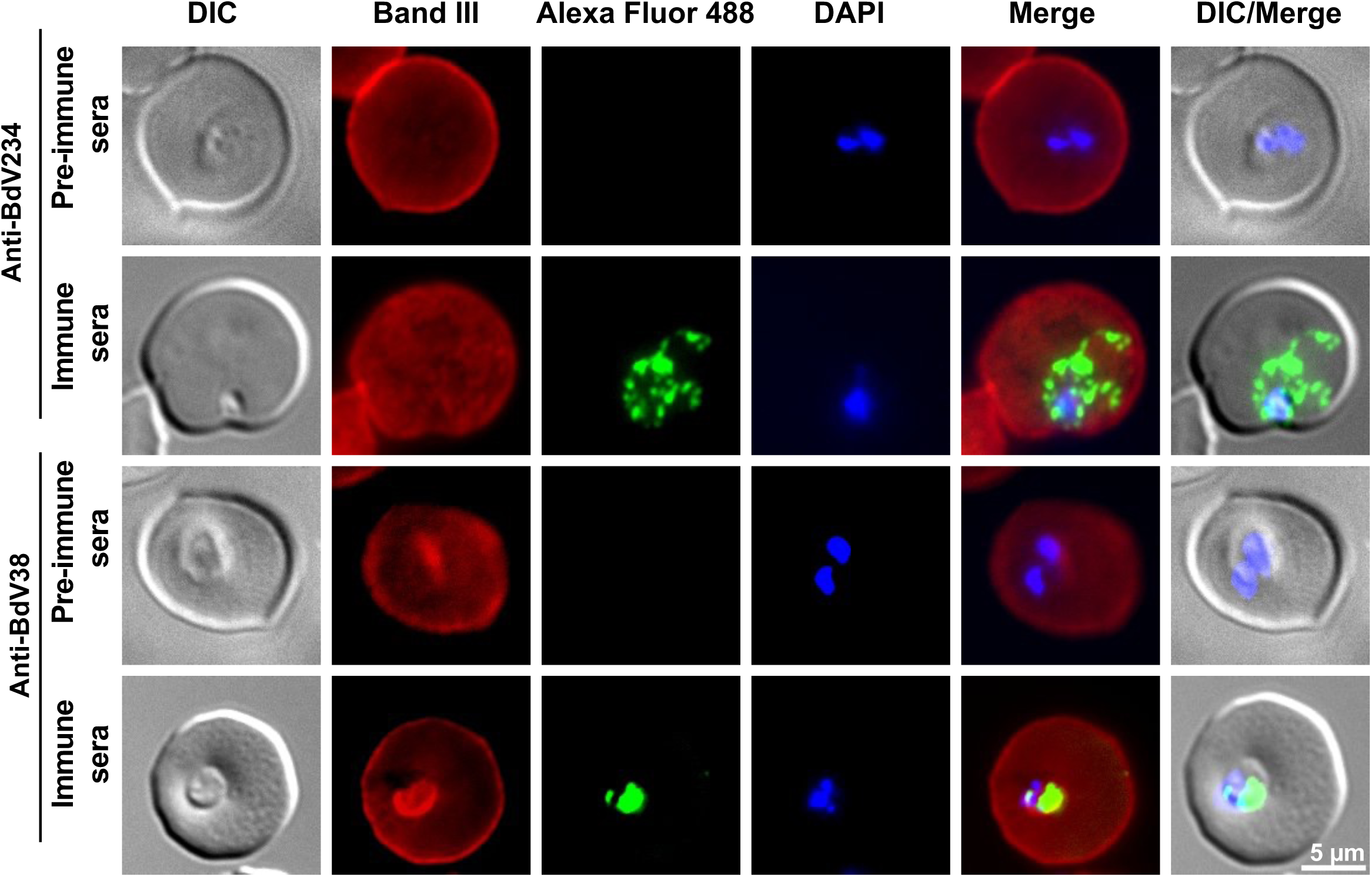
Immunofluorescence assay for the detection of subcellular localization of *B. duncani* secreted proteins. Immunofluorescence staining of BdV234 and BdV38 proteins using rabbit polyclonal anti-BdV234 and BdV38 antibodies. Samples were subsequently, stained with an anti-rabbit secondary antibody conjugated with Alexa Fluor (AF)-488 (Green) in *B. duncani*-infected human RBCs. Anti–Band-III monoclonal antibody (Red) was used to label the human erythrocyte plasma membrane, and DAPI (Blue) was used to stain the parasite nucleus (DNA). The green fluorophore reveals both the localization and distribution of the parasite-secreted proteins within or outside the host red blood cell. Immune Sera: rabbit sera collected after injection of recombinant BdV234 and BdV38 recombinant proteins, and preimmune sera: same rabbit sera collected before injection of recombinant proteins. Differential Interference Contrast (DIC). Scale bars, 5µm.

### Development of *B. duncani* antigen capture Assays BdACAs

The discovery that BdV234 and BdV38 could be readily detected in the hemolysate and environment of *B. duncani*-infected erythrocytes led us to investigate the use of these two proteins as biomarkers of an active *B. duncani* infection. We used the anti-BdV234 and anti-BdV38 antibodies to design an antigen capture sandwich ELISA assay (BdACA) to detect the exported proteins in *B. duncani*-infected blood (**Fig. 3A**). The BdACA assays, one for each antigen, were initially optimized by employing two-fold serial dilutions of recombinant BdV234 and BdV38 proteins to generate a standard curve. The ELISA data was graphed with optical density against known antigen concentrations to obtaining sigmoidal curves (**Fig. 3B**). To further validate the assays for native proteins at the cellular level, an *in vitro* culture of *B. duncani*-infected human erythrocytes at 18% parasitemia with 5% hematocrit was prepared, and the supernatant and hemolysate fractions were collected, subjected to two-fold serial dilutions, and used to detect the exported antigens. From our findings, both BdV234 and BdV38 effectively identified the presence of total parasite proteins that were secreted into the culture supernatant and established a positive correlation with the percentage of parasitemia counted by microscopy (**Fig. 3C**). In addition, when the assays were conducted on the culture hemolysate samples, we observed a comparable correlation accompanied by a marginal rise in the absorbance values (**Fig. 3D**). In both assay conditions, we observed nearly undetectable levels of the proteins when using the supernatant and hemolysates from cultures of uninfected human RBCs. We extended the BdACA validation into the established *in vivo* mouse model, utilizing serum and plasma from *B. duncani*-infected mice (with ∼24% parasitemia) (**Fig. 3E**), along with the H fractions (**Fig. 3F**). We observed robust detection of the parasite antigens when probing the mouse hemolysate fractions with both antibodies, while the levels were merely undetectable in the serum or plasma fractions (**Fig. 3E**). As expected, there was no detection in the serum, plasma, or hemolysate fractions from uninfected mice.

**Figure 3.**
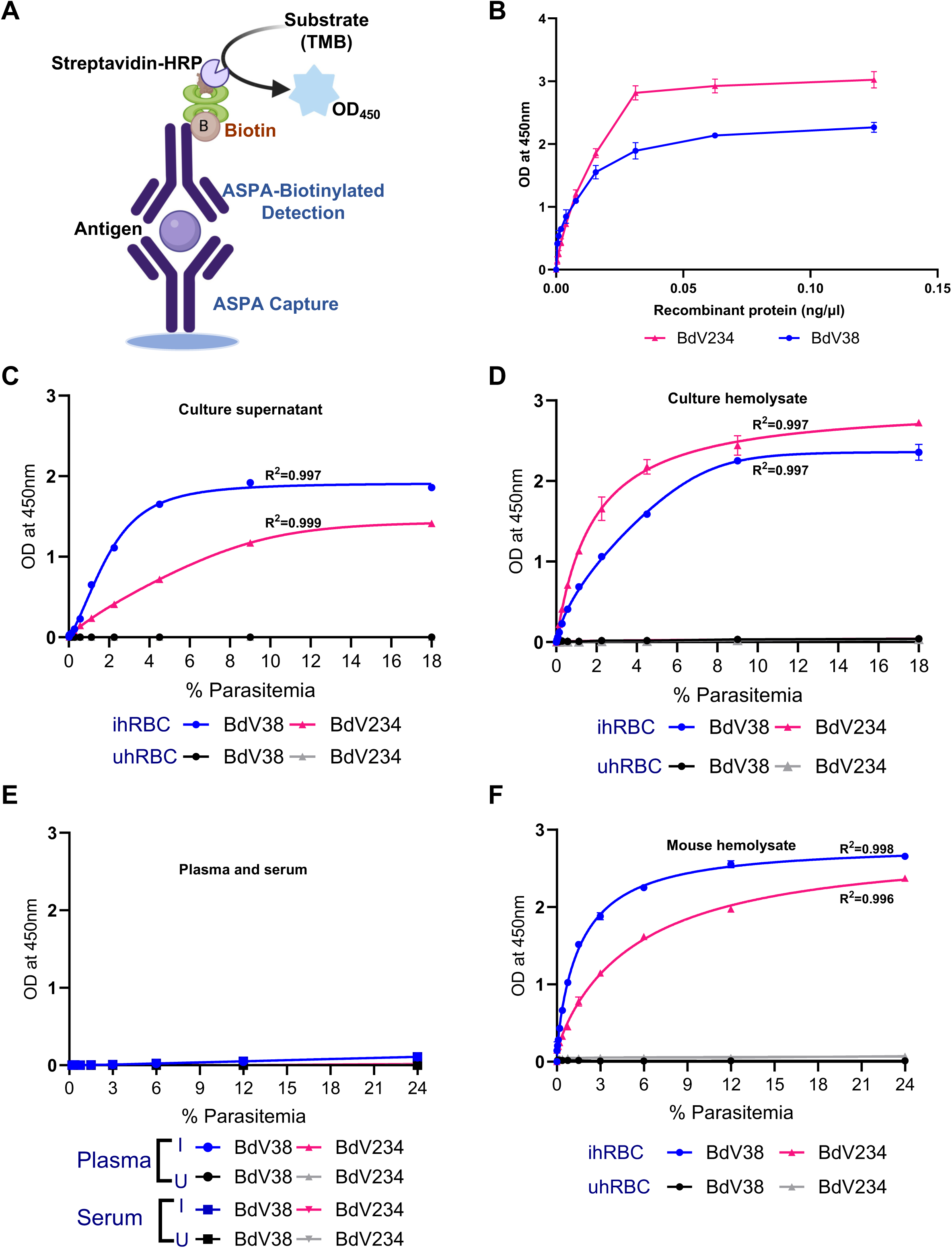
Development and detection of active *B. duncani* infection by BdACA. (**A)** Schematic representation of the ELISA-based *B. duncani* antigen capture assay (BdACA). *B. duncani* secreted antigens (recombinant proteins) BdV234 and BdV38 were used as specific biomarkers for the detection of parasite infection. Respective antigen-specific polyclonal antibodies (ASPA) were used as both capture and biotinylated-detection antibodies to detect the *B. duncani* secreted antigens (BdV234 and BdV38). (**B)** A standard curve with recombinant BdV234 and BdV38 proteins was tested in two-fold serial dilutions. **(C-F)** A dose-response curve showing the absorbance at 450 nm (y-axis) plotted against two-fold serial diluted supernatant **(C)** and hemolysate **(D)** obtained from *in vitro* cultures of *B. duncani* infected human erythrocytes and from *B. duncani* infected mouse plasma/serum **(E)** and hemolysate **(F)** along with uninfected hRBC/mRBC as controls (black and grey). The specific polyclonal antibodies were used to detect BdV234 (pink, triangle) and BdV38 (blue, circle) antigens. The error bars represent the standard deviation (SD), calculated using GraphPad Prism 9.3 software. All samples were analyzed in two independent experiments in triplicates.

### High sensitivity of BdACAs for detection of *B. duncani* infection

The sensitivity of the BdACAs was assessed using a culture of *B. duncani* at ∼9 x 10^6^ *B. duncani*-iRBCs per ml, which was subsequently subjected to two-fold dilutions and allowed to proliferate for 24h in culture. For the comparison of BdACA with qPCR, we isolated the genomic DNA from the same samples and used it for PCR detection of the *B. duncani* 18s rRNA (**Fig. 4A**). The H fractions from the same cultures were further used to evaluate the sensitivity of the BdACAs. The Bd38ACA could detect as low as 1 x 10^3^ iRBCs, while Bd234ACA could detect as low as 2 x 10^3^ iRBCs (**Fig. 4B-C**). No reactivity was observed in uninfected human RBCs, as expected. The limit of blank (LOB) marked the threshold above which all values were considered positive, while the limit of detection (LOD) for Bd38ACA and Bd234ACA was determined to be 3 x of LOB (OD_450_=0.3). The sensitivity of BdACA was also assessed to establish the minimum detectable level of parasitemia. A 1 ml of *B. duncani in vitro* culture with an initial parasitemia of 0.5% (5% hematocrit) was allowed to grow over time, and parasitemia was monitored. After isolating the hemolysate (1 ml), genomic DNA extraction was performed to facilitate quantitative polymerase chain reaction (qPCR) analysis (**Fig. 4D**). The BdACA assay employed 5 microliters per well alongside the previously mentioned hemolysate volume. Our findings demonstrated that the BdACA method effectively identified parasitemia levels as low as 0.5%, consistent with the expected sensitivity. Additionally, as the secretion of antigens increased over time, our assays could detect more secreted antigens (**Fig. 4E-F**). Similarly, to validate the sensitivity of the assay in *vivo* samples, blood was collected from day post-infection (dpi) 0-7 from C3H/HeJ mice. Genomic DNA isolated from parasites at different time points was used in qPCR using *B. duncani* 18s rRNA forward and reverse primers that detected parasites as early as 0-dpi (**Fig. 4G**), while Bd38ACA and Bd234ACA could detect the antigens as early as 2 and 3 dpi, respectively (**Fig. 4H-I**).

**Figure 4.**
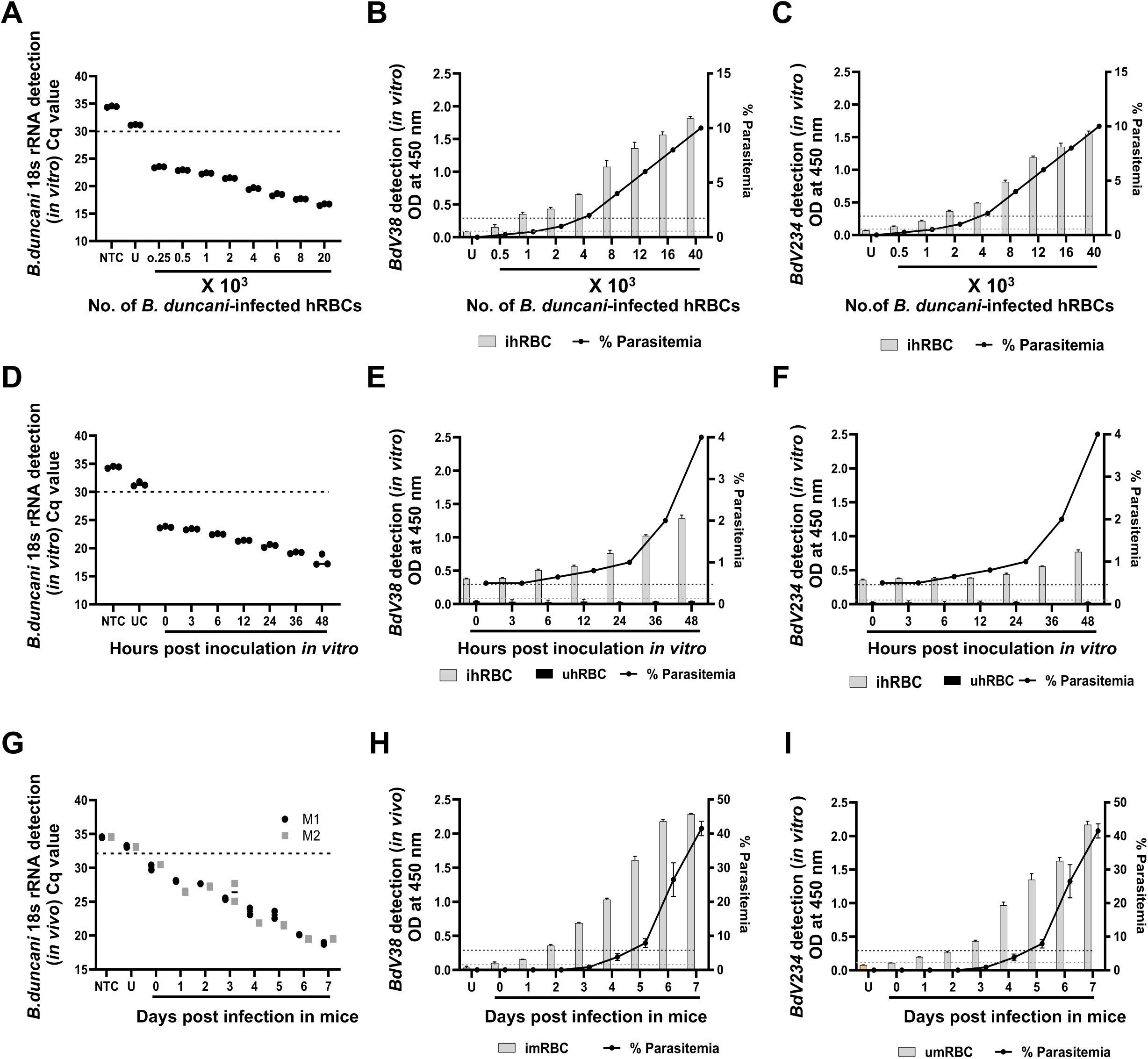
BdACA shows high sensitivity to detect *B. duncani*-infected human and mouse red blood cells. **(A)** Scatter plot illustrates the corelation between cycle threshold values (Cq) and the number of *B. duncani*-infected red blood cells (iRBCs). Cq values were obtained through q-PCR analysis using Bd ITS (18S rRNA) primers on isolated genomic DNA from indicated parasites. The graph ploted using Cq values on the Y-axis against the number of iRBCs on the X-axis, from an *in vitro B. duncani* culture. Horizontal dotted line represents the cut-off threshold, with numerical values on the x-axis depicting the quantity of infected RBCs, where ‘x 10**^3^**’ denotes a scale of 1000 for RBC count. **(B-C)** Detection of BdV38 **(B)** and BdV234 **(C)** secreted antigens from *B. duncani*-infected human RBCs from *in vitro* culture samples with different dilutions of parasites as shown in the bar graph, and the % parasitemia was counted by Giemsa stain (black scatter plot). **(D-F)** *Babesia duncani in vitro* culture, diluted to 0.5% parasitemia (5% hematocrite), was incubated at 37°C. Samples were collected over time, beginning zero hour post-inoculationt to 48 hours. Genomic DNA was isolated from these samples and quantified using qPCR targeting the 18S rRNA gene (**D**). Similarly, hemolysate fractions were prepared to detect the BdV38 (**E**) and BdV234 (**F**) antigens from these samples. **(G-I)** For sensitivity analysis from *in vivo* samples, C3H/HeJ mice were inoculated with 10^6^ parasites and monitored. Parasites were collected at various time points (0-7 dpi) post-infection and subjected for quantification of the 18S rRNA gene by qPCR (**G**). BdACAs detection of BdV38 (**H**) and BdV234 **(I**) antigens from *B. duncani* infected mouse erythrocytes after post-inoculation. Blood from uninfected mice was utsed as control. The dashed lines show the limit of blank (LOB, gray) and the limit of detection (LOD, black), respectively. NTC: non template control, UC: uninfected control, iRBC: infected red blood cells, uRBC: uninfected red blood cells.

### BdV38 and BdV234 as biomarkers for evaluating antibabesial treatment efficacy

An ideal biomarker of active *Babesia* infection is one whose levels show an excellent positive correlation with parasitemia levels and does not remain in the blood following clearance of infection due to drug treatment. To assess whether BdV38 and BdV234 meet these criteria, BdACAs were conducted on *in vitro B. duncani* cultures under both untreated and drug-treated conditions. Parasite cultures with 1 % parasitemia were initially inoculated and collected at 12h intervals over a 72h period (three parasite erythrocytic lifecycles) following treatment with either vehicle control (DMSO) or tafenoquine (TQ) at 3 µM (1x IC_50_) and 6 µM (2x IC_50_). The samples were assessed for parasitemia detection in both conditions through microscopic analysis of Giemsa-stained blood smears. Observations from Giemsa-stained blood smears at stipulated time intervals of post-inoculation displayed continuous parasite growth until 72h (**Fig. 5A**). The decline in % parasitemia over time and the inhibitory impact of 1x and 2x IC_50_ of tafenoquine were evident, detecting parasitemia as low as 0.05% and 0.1% at 48h compared to the untreated (UT) parasites, which reached approximately 10% after 72h post-inoculation (**Fig. 5A)**. The detection of BdV38 from the total parasite lysate positively correlated with the viability of the parasite over time in untreated (UT) samples (**Fig. 5B**). In contrast, the curve declined over time and reached a plateau after 48h with tafenoquine treatment (1x or 2x) (**Fig. 5B**). For post-drug treatment, as tafenoquine cleared the parasites from the culture, the total absorbance showed nearly no difference compared to the uninfected RBC lysate. Upon qPCR analysis of the same samples to assess parasite detection, the presence of *B. duncani* DNA was indicated in both the untreated and treated conditions at stipulated time intervals. However, a clear difference was observed between the uninfected and infected samples (**Fig. 5C**). Similarly, we used whole blood samples from uninfected and *B. duncani*-infected (10^6^) mice that were treated with tafenoquine (TQ) or vehicle. Blood samples were tested to assess parasite growth by microscopy (**Fig. 5D**), and BdV38ACA was used to detect BdV38 secretion that declined after 7 dpi once the parasites were cleared by the drug treatment (**Fig. 5E**). qPCR results suggested DNA remained in circulation even after the parasites were cleared (**Fig. 5F**). Thus, the Bd38ACA assay reliably detected only active infections. Similarly we tested these *in vito* and *in vivo* samples with anti-BdV234 antibody and found the similar results as with BdV38 (Fig. S3).

**Figure 5.**
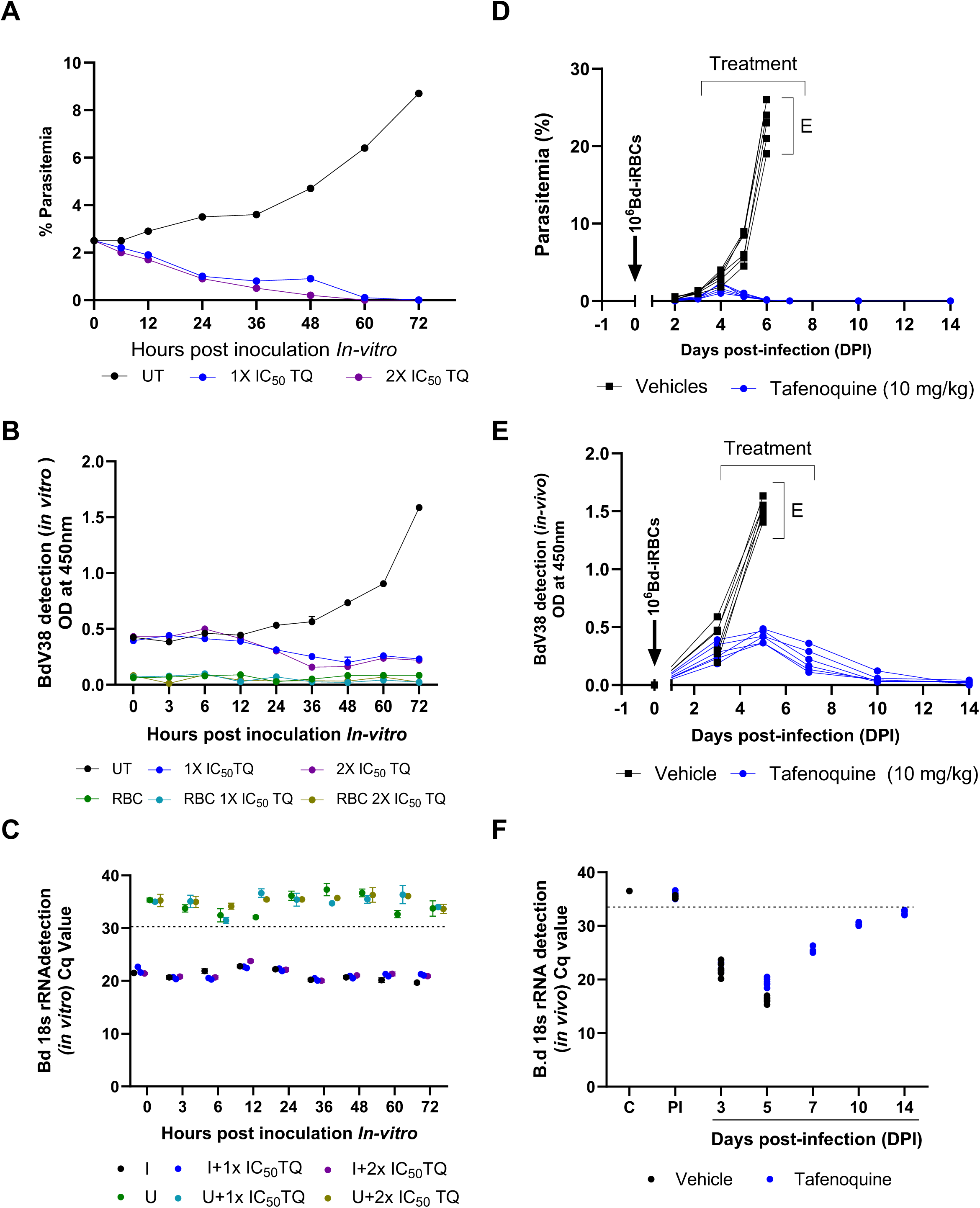
Detection of BdV38 antigen after Tafenoquine treatment of *B. duncani* cultures. **(A and D)** Percent parasitemia over time from *B. duncani in vitro* culture (**A**) or mouse infected with *B. duncani* **(D)** in vehicale (PEG-400) untreated (UT) or tafenoquine treated (1x IC_50_ and 2x IC_50_ TQ samples over time. **(B and E)** Bd38ACA detection of BdV38 antigen in *B. duncani in vitro* culture hemolysate **(B)** and *B. duncani* 10^6^ infected mouse hemolysate **(E**) in untreated or tafenoquine treated samples. **(C and F)** genomic DNA isolated from parasites from *in vitro* **(C)** or mice **(F)** at different time points pre and post tafenoquine treatment were subjected to *B. duncani* 18S rRNA quantitative PCR. Graphs denote the Cq values over time.

### Bd38ACA and Bd234ACA are highly specific assays for *B. duncani* detection

To assess the specificity of the BdACA assays, we tested blood samples infected with other *Babesia* species and similar apicomplexan parasites, such as *P. falciparum* from *in vitro* culture as well as from animal hosts. We also compared the sensitivity of BdACA from uninfected and *B. microti* (10^6^) infected mouse blood. Uninfected and *B. duncani*-infected mouse blood was used as a control. Our *in vivo* results indicated that Bd38ACA (**Fig. 6A**) and Bd234ACA (**Fig. 6C**) are highly specific for *B. duncani*-secreted proteins compared to *B. microti*. Similarly, the assays were conducted on *in vitro* blood samples from different cultures of *B. divergens* (Rouen87 strain), *Babesia MO1*, or *P. falciparum* (3D7 strain), each with 10% parasitemia grown in human erythrocytes. Our result indicated that Bd38ACA (**Fig. 6B**) and Bd234ACA (**Fig. 6D**) only detect the respective proteins secreted by *B. duncani,* and no signal above the lower limit detection of OD 0.3 could be detected in these samples.

**Figure 6.**
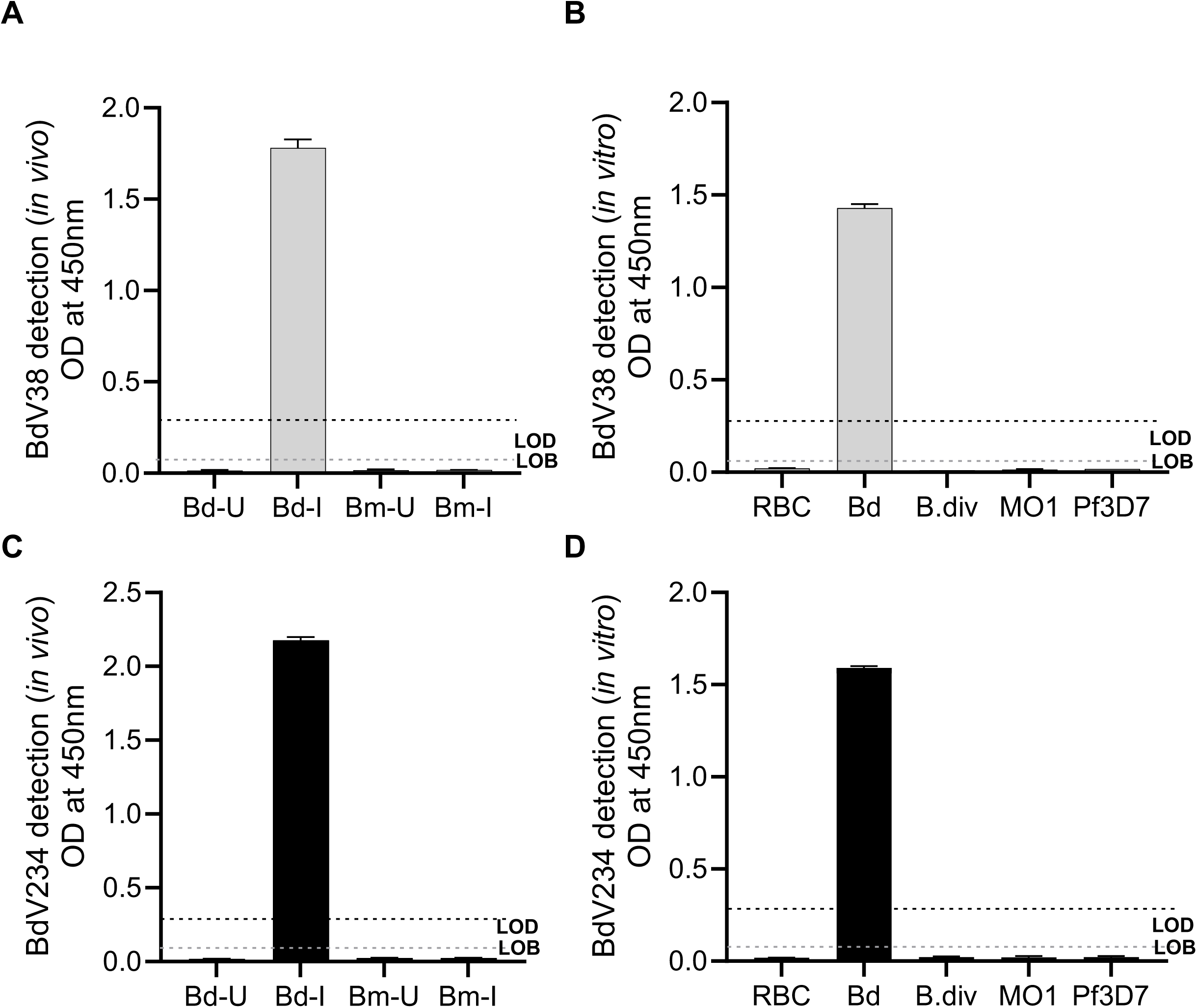
*In vitro* and *In vivo* specificity of BdACAs for *Babesia* antigens. Bd38ACA (**A-B**) or Bd234ACA (**C-D**) were performed on uninfected (Bd-U) or *B. duncani-*infected (Bd-I) (24% parasitemia) C3H/HeJ mouse hemolysate and uninfected (Bm-U) or *B. microti*-infected (Bm-I) (70% parasitemia) CB17 SCID mouse hemolysate (**A and C**). *In vitro* culture hemolysate from uninfected (RBC) or *B. duncani* (Bd) (10 % parasitemia), *B. divergens* Rouen87 (B.div) (10 % parasitemia), *B.* MO1 (10 % parasitemia) (MO1), and *P. falciparum* (*Pf*3D7) (10% parasitemia) infected samples (**B and D**). *Bd*-*Babesia duncani*, *Bm*-*Babesia microti*, *B.div*-*Babesia divergens rouen87*, *Pf*3D7-*Plasmodium falciparum* 3D7, I-infected, U-uninfected, cF12-complete DMEM-F12, and cRPMI-complete RPMI media. The gray dashed lines show the limit of blank (LOB,) and black deshed line show the limit of detection (LOD) (OD_450_=0.3). respectively. Error bars showing the standard deviation (SD) which were calculated by GraphPad Prism 9.3.

### Blood screening using Bd38ACA

To evaluate the effectiveness of BdACA as screening tools for detecting active *B. duncani* infections, we employed the Bd38ACA assay to screen 1,731 whole blood specimens from diverse locations across the US. These encompassed 200 human whole blood samples (100 *B. microti*-positive and 100 *B. microti-*negative) from the American Red Cross (ARC); 770 human whole blood samples from the Lyme Disease Biobank (LDB), previously screened for Lyme disease and *B. microti*; and 761 whole blood samples from white-footed mice collected from Western Pennsylvania **(Fig. 7A**). In this study, the lower limit of detection (LOD; OD_450_ = 0.3) was defined as thrice of the LOB (lower limit of blank), which corresponds to a *B. duncani* parasitemia of 0.3% **(Fig. 7B).** All 1,731 samples, analyzed in this study, showed OD_450_ values below 0.6, which when compared to the standard curve for *B. duncani* parasitemia levels corresponds to parasitemia levels below 0.55% (**Fig. 7A**). Of these 1731 samples, 8 had OD_450_ values between 0.31 and 0.59. Among the 761 white-footed mouse samples, 756 showed signals below the LOD and 5 samples showed signals above the LOD but below an OD_450_ of 0.35 (< than 2x LOD). Similarly, out of the 770 Lyme Disease Biobank samples, 3 showed signals above the LOD with OD_450_ of 0.31, 0.49 and 0.59. The 8 samples with OD_450_ signals above the LOD using the BdV38-based detection assay, were further confirmed to be within this same range when tested using the BdV234ACA assay (not shown), suggesting that these samples either carry *B. duncani* at parasitemia levels below 0.5% or the BdV38 and BdV234 assays detect antigens from other closely related pathogens. To gain further insights into the source of the signals in these samples (N=9, one ARC30 sample also included), PCR-based amplification analyses were conducted using the primer pairs listed in Table 2. Using primers designed to be specific for *B. duncani* (specific to *HSP-70*, *CelTos*, and *BdV38* genes), *B. microti* (specific to *BmITS* and *Bm GPI12* genes), *B. divergens* (specific to AMA-1 gene) and *B. MO1* (specific to the helicase gene), our PCR analyses showed that these samples do not contain DNA from these species (**Fig. S4-S5**). To assess whether these samples contain DNA from other *Babesia* species, we selected samples with the highest OD_450_ among these for PCR analysis using a previously reported primer pair (39) that amplifies a 200 bp of the 18 S rRNA gene from multiple *Babesia* species (**Fig. 6A**). The nine samples included 1 from ARC, 3 from Lyme Biobank and 5 from field mice. As shown in **Fig. 6 B**, six of the 9 samples tested positive by PCR. Sequencing of the 200 bp PCR product revealed sequences with 98 to 100% identity to 18S rRNA genes from *Babesia* species of Clade VI (*B. divergens, capreoli, B. odocoilei,* and *B. venatorum*) **(Table 3).** To further evaluate these data, a new primer set was designed to bind to conserved regions of the 18S rRNA genes of *Babesia* species and amplify a 736 bp. Using this primer set against all 9 samples, we found 5 (field mice) samples to be positive by PCR. However, sequencing of the fragments failed to identify a specific species at more than 90% sequence identify (**Fig. S6 C-D**). Together these data suggest that the 8 samples may contain new species of *Babesia* or closely related organisms that the BdACAs can also detect. However, the exact identity of these new species could not be determined due to limited amount of material available for large scale genomic analyses.

**Figure 7.**
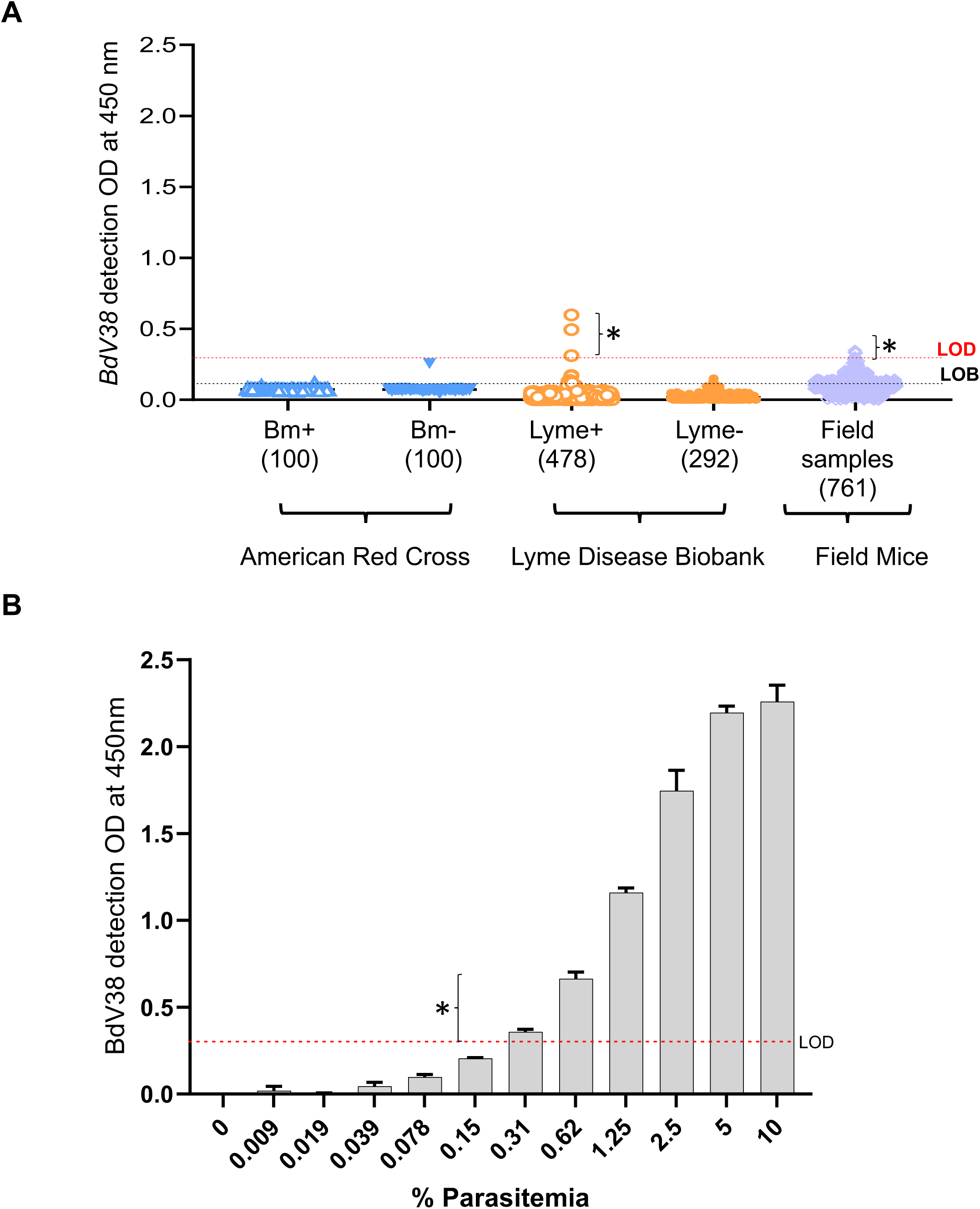
Bd38ACA screening of ARC, LDB human patient samples, and untested field-derived white-footed mouse samples. **(A)** Screening of known positive and negative *Babesia* patient samples, Lyme positive and negative samples, and unknown infection from field-derived white-footed mouse samples with Bd38ACA. One hundred *B. microti* transcription-mediated amplification (TMA)-positive human patient samples (blue open upward triangle), 100 TMA-negative human patient samples (blue closed inverted triangle) from American Red Cross (ARC), 478 Lyme disease positive (orange open circle), and 292 control samples (orange closed circle) from Lyme Disease Biobank (LDB) samples (770 samples) and 761 unknown field mice samples (purple open diamond) were tested. Each sample was used in triplicates. **(B)** Two fold serially dilutaed *B. duncani* infected human RBC hemolysate (ihRBC; gray bar graph) were used as positive controls to compare the samples OD_450_ in corelation with % parasitemia. Black and red dash lines show the limit of blank (LOB) and limit of detection (LOD) respectively. Error bars showing the standard deviation (SD) were calculated using GraphPad Prism 9.3 software. (*) showing OD450 of patient samples that were slightly positive for Bd38ACA.

**Table 2.**
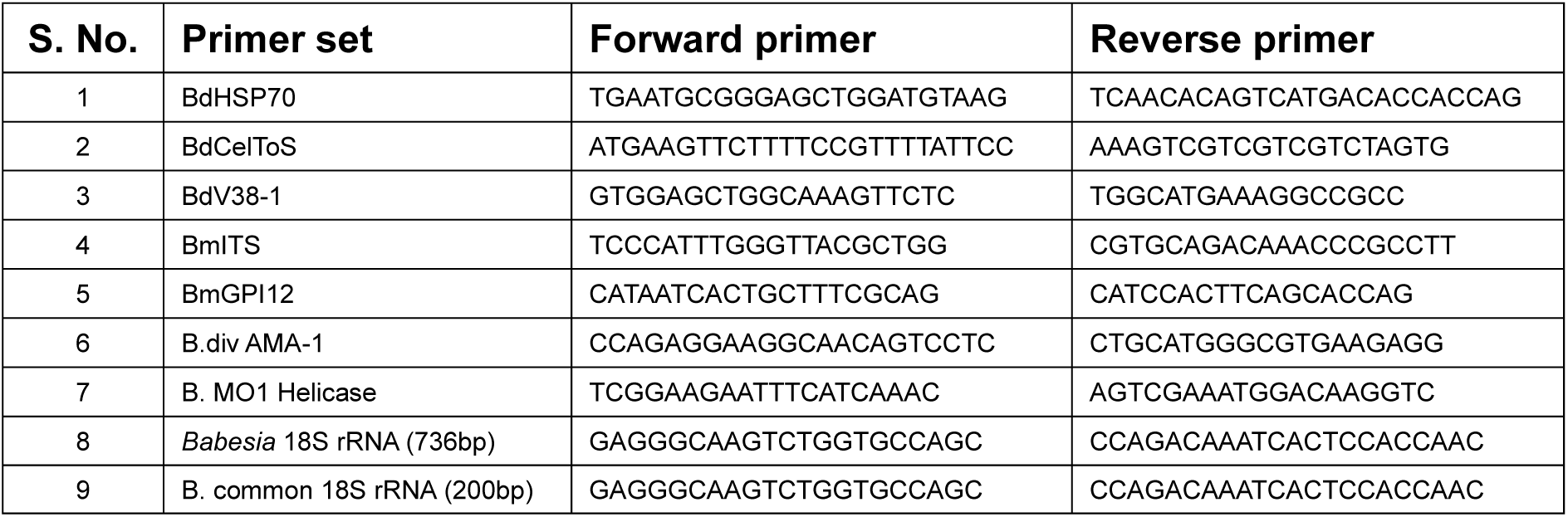
List of primer sequences for *Babesia* and species-specific PCRs.

**Table 3.** List of PCR positive samples with identity scores to 18S rRNA of *Babesia* species genes from of Clade VI.

| Sample ID | Gene description | Species | Percent Identity | Accession No. |
| --- | --- | --- | --- | --- |
| LDB85 | Small subunit ribosomal RNA gene (18S rRNA) | *Babesia odocoilei* isolate JC34 | 99.21 | KY805843.1 |
|  |  | *Babesia odocoilei* isolate JC09 | 99.21 | KY805842.1 |
|  |  | *Babesia capreoli* isolate GM19 | 99.21 | KY805834.1 |
|  |  | *Babesia sp. venatorum* isolate RD8 | 99.21 | MG344777.1 |
| LDB139 | Small subunit ribosomal RNA gene (18S rRNA) | *Babesia odocoilei* isolate JC34 | 98.52 | KY805843.1 |
|  |  | *Babesia odocoilei* isolate JC09 | 98.52 | KY805842.1 |
|  |  | *Babesia capreoli* isolate GM19 | 98.52 | KY805834.1 |
|  |  | *Babesia sp. venatorum* isolate RD8 | 98.52 | MG344777.1 |
| FM1 | Small subunit ribosomal RNA gene (18S rRNA) | *Babesia odocoilei* isolate JC34 | 98.04 | KY805843.1 |
|  |  | *Babesia odocoilei* isolate JC09 | 98.04 | KY805842.1 |
|  |  | *Babesia odocoilei* isolate YB73 | 98.04 | KY805835.1 |
|  |  | *Babesia sp. venatorum* isolate R30 | 98.04 | MG062781.1 |
| FM7 | Small subunit ribosomal RNA gene (18S rRNA) | *Babesia odocoilei* isolate JC34 | 100.00 | KY805843.1 |
|  |  | *Babesia odocoilei* isolate JC09 | 100.00 | KY805842.1 |
|  |  | *Babesia capreoli* isolate GM19 s | 100.00 | KY805834.1 |
|  |  | *Babesia sp. venatorum* isolate RD8 | 100.00 | MG344777.1 |
| FM8 | Small subunit ribosomal RNA gene (18S rRNA) | *Babesia odocoilei* isolate JC34 | 100.00 | KY805843.1 |
|  |  | *Babesia odocoilei* isolate JC09 | 100.00 | KY805842.1 |
|  |  | *Babesia capreoli* isolate GM19 | 100.00 | KY805834.1 |
|  |  | *Babesia sp. venatorum* isolate R30 | 100.00 | MG062781.1 |
| FM15 | Small subunit ribosomal RNA gene (18S rRNA) | *Babesia odocoilei* isolate JC34 | 98.33 | KY805843.1 |
|  |  | *Babesia odocoilei* isolate JC09 | 98.33 | KY805842.1 |
|  |  | *Babesia capreoli* isolate GM19 s | 98.33 | KY805834.1 |
|  |  | *Babesia sp. venatorum* isolate RD8 | 98.33 | MG344777.1 |

## DISCUSSION

Herein, we report the development of the first antigen capture assays, BdACAs, for the detection of active *B. duncani* infection. The assays detect the presence of *B. duncani*-secreted antigens BdV38 (31 kDa) or BdV234 (27 kDa) in blood samples from infected cultures or animals with high sensitivity and specificity. The BdACAs exhibit a remarkable sensitivity in detecting these secreted *B. duncani* antigens from as little as a single drop of blood, equivalent to 1.6 x 10^7^ red blood cells, in both human and mouse samples. Furthermore, the assay specifically distinguishes *B. duncani* infections when tested against other human babesiosis-causing parasites (*B. microti, B. divergens* (Rouen87), *Babesia* MO1) or related protozoan species (*P. falciparum*) that causes human malaria. These assays, therefore, open up new possibilities for efficient and convenient diagnosis of *B. duncani* infection, ensuring timely intervention and improved healthcare outcomes. The versatility of this assay makes it useful in various setups, including point-of-care (POC) tests in clinics or at home. Its potential applications include confirming acute *B. duncani* infection, screening clinical samples, and ensuring the safety of blood supplies at blood donation centers, thereby preventing transfusion-transmitted babesiosis. Additionally, it offers a valuable tool for conducting epidemiological surveys to detect *B. duncani* infection in humans and reservoir hosts. Given the urgent need for improved diagnostic tests to combat vector borne or transfusion-transmitted babesiosis, as seen with other *Babesia sp.* in the USA, the development of this assay represents a significant stride toward achieving that goal (40, 41) and the first report of an ELISA based antigen capture assay for *B. duncani*.

The current strategies employed for the detection of subclinical infections of intraerythrocytic parasites suffer from notable limitations. Microscopy, although widely used, exhibits low sensitivity and specificity, making it challenging to differentiate between various species of *Babesia* and other apicomplexan parasites, such as *Plasmodium*. Serological tests such as immunofluorescence assays (IFA) lack reliability in distinguishing between acute and chronic infections. On the other hand, PCR based assays that show high sensitivity to detect parasite DNA can detect the DNA even after the infection has been cleared, as observed in the case of *B. microti* (22). An ineffective diagnostic tool also hampers the proper understanding of the epidemiological distribution of the parasite in its reservoir hosts.

One key advantage leveraged in our approach is the use of an in culture-in mouse (ICIM) model system of *B. duncani* (42) infection, which allowed us to systematically examine different aspects of our assay, including optimizing the assay using various sample types such as hemolysate, supernatant, whole blood, plasma, and serum. Additionally, the recent comprehensive genome analysis of *B. duncani* by Singh *et. al.* (29) shed light on the abundance and nature of proteins, epigenome, and transcriptome analysis identified classes of candidate virulence factors, antigens for diagnosis of active infection, and several attractive drug targets. Studies in *B. microti* have shown that secreted proteins are excellent biomarkers for the detection of active parasite infection (43, 44). These findings led to the development of antigen capture assays (ACAs) for the detection of the secreted antigen BmGPI12 (22, 23). Here, we have used an integrated approach consisting of genomic, transcriptomic, and proteomic analyses to identify secreted proteins of *B. duncani,* which could be suitable as biomarkers of active infection from different fractions (S, H, and P) **(Table 1)**. Among the pool of exported proteins, BdV234 and BdV38 were specifically chosen due to their substantial peptide abundance and notably elevated expression levels **(Table 1)**. We reasoned that those proteins with higher abundance and containing signal sequences were predominantly destined for the extracellular space, where they could play a crucial role in eliciting cell signaling and facilitating communication between host and parasite to promote infection propagation. To our surprise, our initial attempts to detect BdV234 and BdV38 directly from the plasma or serum of *B. duncani*-infected mice were unsuccessful. However, by employing ultracentrifugation and the Exo-Quick approach, we were able to purify extracellular vesicles (EVs) that contained these parasite proteins. The EVs containing these *B. duncani* proteins may play a crucial role in the trafficking and delivery of these proteins, allowing them to be expressed on the outer membrane of infected RBCs similar to that reported for *B. microti* (30, 38). Our IFA results, from *in vitro B. duncani-*infected parasites, further support the cellular distribution of these proteins. Further investigation is warranted to explore the precise mechanism by which these proteins are sorted and packaged into EVs, as well as their specific role in mediating their transport to the surface of infected RBCs. Understanding the molecular processes involved in this vesicular transport machinery will contribute to our overall comprehension of *B. duncani* pathogenesis and host-parasite interactions.

This study assessed the field suitability of Bd38ACA by screening a diverse array of human and mouse whole blood samples. The assay exhibited minimal cross-reactivity with both human and field-derived white-footed mouse samples. However, a subset (0.4%) of samples showed low reactivity with Bd38ACA, at levels significantly lower in comparison to the positive control samples. Subsequent PCR analysis confirmed the absence of *B. duncani* parasites in these samples but the exact organisms, likely a species closely related to *B. duncani*, that cause the cross-reactivity detected in the BdACA assays remain to be further characterized.

In conclusion, our findings demonstrate that BdACA is a reliable diagnostic assay for the detection of active *B. duncani* infection. It is suitable for large-scale screening of the blood supply and further optimization of the assay is warranted to test uninfected and infected human blood samples and compare with the current method (NAT assays) for blood screening. The development of such a diagnostic tool is a significant step forward in the fight against *B. duncani* and can significantly improve the accuracy and efficiency of diagnosis, treatment, and surveillance of this emerging tick-borne disease.

## Data Availability

All data produced in the present work are contained in the manuscript

## Author contributions

MC, PV, DD, LZ, JC, and AP: Investigation, methodology, formal analysis, visualization, writing original draft, review, and editing. SW: Sample collection and analysis. CB: Conceptualization, supervision, funding acquisition, project administration, writing original draft, review, and editing. All authors have read and agreed to the published version of the manuscript. All authors contributed to the article and approved the submitted version.

## Funding

The research described herein was supported by the Global Lyme Alliance Foundation. CBM research is also supported by NIH grants AI138139, AI123321, AI152220 and AI153100, and AI136118, the Steven and Alexandra Cohen Foundation (Lyme 62 2020), and The Blavatnik Family Foundation.

## Acknowledgments

We thank the American Red Cross and the Lyme Disease Biobank for providing the blood samples used in this study.

## Notes

### Competing Interest Statement

The authors have declared no competing interest.

### Author Declarations

The use of de-identified human blood samples in this study was following an approved Yale IRB protocol to Dr. Ben Mamoun.

## References

1. Jones KE, Patel NG, Levy MA, Storeygard A, Balk D, Gittleman JL, Daszak P. 2008. Global trends in emerging infectious diseases. Nature 451:990–3.

2. Eisen RJ, Eisen L, Beard CB. 2016. County-Scale Distribution of Ixodes scapularis and Ixodes pacificus (Acari: Ixodidae) in the Continental United States. J Med Entomol 53:349–86.

3. Eisen RJ, Kugeler KJ, Eisen L, Beard CB, Paddock CD. 2017. Tick-Borne Zoonoses in the United States: Persistent and Emerging Threats to Human Health. Ilar j 58:319–335.

4. Rosenberg R, Lindsey NP, Fischer M, Gregory CJ, Hinckley AF, Mead PS, Paz-Bailey G, Waterman SH, Drexler NA, Kersh GJ, Hooks H, Partridge SK, Visser SN, Beard CB, Petersen LR. 2018. Vital Signs: Trends in Reported Vectorborne Disease Cases - United States and Territories, 2004-2016. MMWR Morb Mortal Wkly Rep 67:496–501.

5. Renard I, Ben Mamoun C. 2021. Treatment of Human Babesiosis: Then and Now. Pathogens 10.

6. Swanson M PA, Williamson J, Montgomery S. 2023. Trends in Reported Babesiosis Cases — United States, 2011–2019.

7. Puri A, Bajpai S, Meredith S, Aravind L, Krause PJ, Kumar S. 2021. Babesia microti: Pathogen Genomics, Genetic Variability, Immunodominant Antigens, and Pathogenesis. Front Microbiol 12:697669.

8. Vannier E, Gewurz BE, Krause PJ. 2008. Human babesiosis. Infect Dis Clin North Am 22:469–88, viii-ix.

9. Vannier EG, Diuk-Wasser MA, Ben Mamoun C, Krause PJ. 2015. Babesiosis. Infect Dis Clin North Am 29:357–70.

10. Gray EB HB. 2019. Babesiosis Surveillance — United States, 2011–2015. MMWR Surveill Summ 68:1–11.

11. O’Connor KE, Kjemtrup AM, Conrad PA, Swei A. 2018. An Improved PCR Protocol For Detection of Babesia duncanI In Wildlife and Vector Samples. J Parasitol 104:429–432.

12. Conrad PA, Kjemtrup AM, Carreno RA, Thomford J, Wainwright K, Eberhard M, Quick R, Telford SR, 3rd, Herwaldt BL. 2006. Description of Babesia duncani n.sp. (Apicomplexa: Babesiidae) from humans and its differentiation from other piroplasms. Int J Parasitol 36:779–89.

13. Swei A, O’Connor KE, Couper LI, Thekkiniath J, Conrad PA, Padgett KA, Burns J, Yoshimizu MH, Gonzales B, Munk B, Shirkey N, Konde L, Ben Mamoun C, Lane RS, Kjemtrup A. 2019. Evidence for transmission of the zoonotic apicomplexan parasite Babesia duncani by the tick Dermacentor albipictus. Int J Parasitol 49:95–103.

14. Quick RE, Herwaldt BL, Thomford JW, Garnett ME, Eberhard ML, Wilson M, Spach DH, Dickerson JW, Telford SR, 3rd, Steingart KR, Pollock R, Persing DH, Kobayashi JM, Juranek DD, Conrad PA. 1993. Babesiosis in Washington State: a new species of Babesia? Ann Intern Med 119:284–90.

15. Herwaldt BL, Kjemtrup AM, Conrad PA, Barnes RC, Wilson M, McCarthy MG, Sayers MH, Eberhard ML. 1997. Transfusion-transmitted babesiosis in Washington State: first reported case caused by a WA1-type parasite. J Infect Dis 175:1259–62.

16. Bloch EM, Herwaldt BL, Leiby DA, Shaieb A, Herron RM, Chervenak M, Reed W, Hunter R, Ryals R, Hagar W, Xayavong MV, Slemenda SB, Pieniazek NJ, Wilkins PP, Kjemtrup AM. 2012. The third described case of transfusion-transmitted Babesia duncani. Transfusion 52:1517–22.

17. Kjemtrup AM, Conrad PA. 2000. Human babesiosis: an emerging tick-borne disease. Int J Parasitol 30:1323–37.

18. Fritz CL, Kjemtrup AM, Conrad PA, Flores GR, Campbell GL, Schriefer ME, Gallo D, Vugia DJ. 1997. Seroepidemiology of emerging tickborne infectious diseases in a Northern California community. J Infect Dis 175:1432–9.

19. Persing DH, Herwaldt BL, Glaser C, Lane RS, Thomford JW, Mathiesen D, Krause PJ, Phillip DF, Conrad PA. 1995. Infection with a babesia-like organism in northern California. N Engl J Med 332:298–303.

20. Scott JD, Scott CM. 2018. Human Babesiosis Caused by Babesia duncani Has Widespread Distribution across Canada. Healthcare (Basel) 6.

21. Healy GR, Speilman A, Gleason N. 1976. Human babesiosis: reservoir in infection on Nantucket Island. Science 192:479–80.

22. Thekkiniath J, Mootien S, Lawres L, Perrin BA, Gewirtz M, Krause PJ, Williams S, Doggett JS, Ledizet M, Ben Mamoun C. 2018. BmGPAC, an Antigen Capture Assay for Detection of Active Babesia microti Infection. J Clin Microbiol 56.

23. Gagnon J, Timalsina S, Choi JY, Chand M, Singh P, Lamba P, Gaur G, Pal AC, Mootien S, Marcos LA, Ben Mamoun C, Ledizet M. 2022. Specific and Sensitive Diagnosis of Babesia microti Active Infection Using Monoclonal Antibodies to the Immunodominant Antigen BmGPI12. J Clin Microbiol 60:e0092522.

24. Caminade C, McIntyre KM, Jones AE. 2019. Impact of recent and future climate change on vector-borne diseases. Ann N Y Acad Sci 1436:157–173.

25. Gilbert L. 2021. The Impacts of Climate Change on Ticks and Tick-Borne Disease Risk. Annu Rev Entomol 66:373–388.

26. Abraham A, Brasov I, Thekkiniath J, Kilian N, Lawres L, Gao R, DeBus K, He L, Yu X, Zhu G, Graham MM, Liu X, Molestina R, Ben Mamoun C. 2018. Establishment of a continuous in vitro culture of Babesia duncani in human erythrocytes reveals unusually high tolerance to recommended therapies. J Biol Chem 293:19974–19981.

27. Kumari V, Pal AC, Singh P, Mamoun CB. 2022. Babesia duncani in Culture and in Mouse (ICIM) Model for the Advancement of Babesia Biology, Pathogenesis, and Therapy. Bio Protoc 12.

28. Singh P, Pal AC, Mamoun CB. 2022. An Alternative Culture Medium for Continuous In Vitro Propagation of the Human Pathogen Babesia duncani in Human Erythrocytes. Pathogens 11.

29. Singh P, Lonardi S, Liang Q, Vydyam P, Khabirova E, Fang T, Gihaz S, Thekkiniath J, Munshi M, Abel S, Ciampossin L, Batugedara G, Gupta M, Lu XM, Lenz T, Chakravarty S, Cornillot E, Hu Y, Ma W, Gonzalez LM, Sánchez S, Estrada K, Sánchez-Flores A, Montero E, Harb OS, Le Roch KG, Mamoun CB. 2023. Babesia duncani multi-omics identifies virulence factors and drug targets. Nat Microbiol 8:845–859.

30. Thekkiniath J, Kilian N, Lawres L, Gewirtz MA, Graham MM, Liu X, Ledizet M, Ben Mamoun C. 2019. Evidence for vesicle-mediated antigen export by the human pathogen Babesia microti. Life Sci Alliance 2.

31. Baranyai T, Herczeg K, Onódi Z, Voszka I, Módos K, Marton N, Nagy G, Mäger I, Wood MJ, El Andaloussi S, Pálinkás Z, Kumar V, Nagy P, Kittel Á, Buzás EI, Ferdinandy P, Giricz Z. 2015. Isolation of Exosomes from Blood Plasma: Qualitative and Quantitative Comparison of Ultracentrifugation and Size Exclusion Chromatography Methods. PLoS One 10:e0145686.

32. Chand M, Choi JY, Pal AC, Singh P, Kumari V, Thekkiniath J, Gagnon J, Timalsina S, Gaur G, Williams S, Ledizet M, Mamoun CB. 2022. Epitope profiling of monoclonal antibodies to the immunodominant antigen BmGPI12 of the human pathogen Babesia microti. Front Cell Infect Microbiol 12:1039197.

33. Wang Y, Zhang S, Wang J, Rashid M, Wang X, Liu X, Yin H, Guan G. 2022. Nested qPCR assay to detect Babesia duncani infection in hamsters and humans. Parasitol Res 121:3603–3610.

34. Wang G, Wormser GP, Zhuge J, Villafuerte P, Ip D, Zeren C, Fallon JT. 2015. Utilization of a real-time PCR assay for diagnosis of Babesia microti infection in clinical practice. Ticks Tick Borne Dis 6:376–82.

35. Horn EJ, Dempsey G, Schotthoefer AM, Prisco UL, McArdle M, Gervasi SS, Golightly M, De Luca C, Evans M, Pritt BS, Theel ES, Iyer R, Liveris D, Wang G, Goldstein D, Schwartz I. 2020. The Lyme Disease Biobank: Characterization of 550 Patient and Control Samples from the East Coast and Upper Midwest of the United States. J Clin Microbiol 58.

36. Tamburro D, Fredolini C, Espina V, Douglas TA, Ranganathan A, Ilag L, Zhou W, Russo P, Espina BH, Muto G, Petricoin EF, 3rd, Liotta LA, Luchini A. 2011. Multifunctional core-shell nanoparticles: discovery of previously invisible biomarkers. J Am Chem Soc 133:19178–88.

37. Magni R, Luchini A, Liotta L, Molestina RE. 2019. Analysis of the Babesia microti proteome in infected red blood cells by a combination of nanotechnology and mass spectrometry. Int J Parasitol 49:139–144.

38. Beri D, Rodriguez M, Singh M, Liu Y, Rasquinha G, An X, Yazdanbakhsh K, Lobo CA. 2022. Identification and characterization of extracellular vesicles from red cells infected with Babesia divergens and Babesia microti. Front Cell Infect Microbiol 12:962944.

39. Qurollo BA, Archer NR, Schreeg ME, Marr HS, Birkenheuer AJ, Haney KN, Thomas BS, Breitschwerdt EB. 2017. Improved molecular detection of Babesia infections in animals using a novel quantitative real-time PCR diagnostic assay targeting mitochondrial DNA. Parasit Vectors 10:128.

40. Herwaldt BL, Linden JV, Bosserman E, Young C, Olkowska D, Wilson M. 2011. Transfusion-associated babesiosis in the United States: a description of cases. Ann Intern Med 155:509–19.

41. Fang DC, McCullough J. 2016. Transfusion-Transmitted Babesia microti. Transfus Med Rev 30:132–8.

42. Pal AC, Renard I, Singh P, Vydyam P, Chiu JE, Pou S, Winter RW, Dodean R, Frueh L, Nilsen AC, Riscoe MK, Doggett JS, Ben Mamoun C. 2022. Babesia duncani as a Model Organism to Study the Development, Virulence, and Drug Susceptibility of Intraerythrocytic Parasites In Vitro and In Vivo. J Infect Dis 226:1267–1275.

43. Elton CM, Rodriguez M, Ben Mamoun C, Lobo CA, Wright GJ. 2019. A library of recombinant Babesia microti cell surface and secreted proteins for diagnostics discovery and reverse vaccinology. Int J Parasitol 49:115–125.

44. Luo Y, Jia H, Terkawi MA, Goo YK, Kawano S, Ooka H, Li Y, Yu L, Cao S, Yamagishi J, Fujisaki K, Nishikawa Y, Saito-Ito A, Igarashi I, Xuan X. 2011. Identification and characterization of a novel secreted antigen 1 of Babesia microti and evaluation of its potential use in enzyme-linked immunosorbent assay and immunochromatographic test. Parasitol Int 60:119–25.

